# Machine learning-based advanced coronary artery disease pretest probability model: Comparison with conventional pretest probability models

**DOI:** 10.64898/2026.03.25.26348861

**Authors:** Youngtaek Hong, Jina Lee, Hyung-Bok Park, Wonse Kim, Yeonyee E. Yoon, Hyunseok Jeong, Gaeun Kim, Byungchang So, Junseok Lee, Mayank Dalakoti, Ji Min Sung, Woong Kook, Hyuk-Jae Chang

## Abstract

**Background:** Pretest probability (PTP) models using clinical risk factors guide decision-making for coronary artery disease (CAD). Existing models (Updated Diamond–Forrester [UDF] and CAD Consortium [CAD2]) exhibit suboptimal predictive efficacy in Asian populations due to ethnic differences in atherosclerosis and risk profiles. We developed an advanced CAD-specific PTP model using ridge-penalized logistic regression and validated its reliability.

**Methods:** Utilizing data from 4,696 Korean patients (3 trials and 2 cohorts), we employed ridge regression to develop an advanced PTP model (K-CAD) for identifying patients with CAD with ≥50% diameter stenosis, determined using coronary computed tomography or invasive coronary angiography. External validation used datasets from another tertiary center (External Validation Cohort 1, n=428) and a nationwide health checkup cohort (External Validation Cohort 2, n=117,294). We compared K-CAD with existing models using continuous receiver operating characteristic (ROC) and ternary net reclassification improvement (NRI) analyses.

**Findings:** Continuous ROC analysis in External Validation Cohort 1 revealed areas under the curves (AUCs) for UDF, 0·68 (95% confidence interval [CI] 0·63–0·73); CAD2, 0·71 (95%CI 0·67–0·76), and K-CAD, 0·76 (95%CI 0·71–0·80). K-CAD significantly outperformed UDF (p <0·001) and CAD2 (p <0·05). NRI analysis demonstrated that K-CAD improved reclassification of non-obstructive patients into low-risk categories. External validation using the nationwide dataset (surrogate endpoint: ICD-10 I20) yielded AUCs for UDF, 0·61 (95% CI 0·58–0·64); CAD2, 0·66 (95%CI 0·63–0·69); and K-CAD, 0·67 (95%CI 0·64–0·70).

**Interpretation:** The study demonstrated K-CAD’s utility employing extensive high-quality datasets, highlighting its potential for predicting CAD risk in the Korean population.

**Funding:** This work was supported by a Korean Medical Device Development Fund grant funded by the Korean government (Ministry of Science and ICT; Ministry of Trade, Industry, and Energy; Ministry of Health & Welfare; and Ministry of Food and Drug Safety) (Project Number: 1711139017, RS-2020-KD000156). The funding agency had no role in the study design, data collection, data analysis, data interpretation, or writing of the manuscript.

**Trial registration:** ClinicalTrials.gov Identifier: NCT02803411.

**Research in context:** *Evidence before this study:* We searched PubMed, MEDLINE, and Google Scholar for studies published from January 1, 2000, to December 31, 2024, using the search terms “pretest probability,” “coronary artery disease,” “prediction model,” “Asian,” “Korean,” “machine learning,” and “risk stratification,” without language restrictions. The Updated Diamond–Forrester (UDF) and CAD Consortium (CAD2) clinical models are the most widely endorsed pretest probability (PTP) tools in current American Heart Association/American College of Cardiology and European Society of Cardiology guidelines. However, these models were developed predominantly using Western population data. Studies evaluating their performance in Korean and other East Asian populations consistently reported suboptimal discriminatory ability, with areas under the receiver operating characteristic curve (AUCs) ranging from 0·69 to 0·74. Evidence suggested that ethnic differences in coronary atherosclerosis patterns and cardiovascular risk factor profiles contribute to poor calibration of Western-derived models in Asian cohorts.

*Added value of this study:* This study developed K-CAD, a ridge-penalized logistic regression model specifically calibrated for the Korean population, using a large-scale training dataset of 4,696 patients from three randomized controlled trials and two registries. Unlike existing models that rely solely on age, sex, symptoms, and basic medical history, K-CAD integrates readily available routine laboratory results, including lipid profiles, creatinine, and glycated hemoglobin, into the prediction framework. K-CAD achieved an AUC of 0·76 in an independent external validation cohort of 428 high-risk patients evaluated by invasive coronary angiography, significantly outperforming both UDF (AUC 0·68, p<0·001) and CAD2 (AUC 0·71, p<0·05). Importantly, K-CAD substantially improved risk stratification by reclassifying 79·9% of non-obstructive patients misclassified as high-risk by UDF into lower-risk categories, potentially reducing unnecessary downstream testing. The model’s generalizability was further demonstrated in a nationwide health screening cohort of 117,294 individuals. Additionally, the complete model parameters and an online calculator are publicly available, ensuring transparency and reproducibility.

*Implications of all the available evidence:* The available evidence indicates that Western-derived PTP models systematically overestimate coronary artery disease risk in Korean and East Asian populations, leading to the overclassification of patients into high-risk categories and potentially unnecessary invasive testing. The K-CAD model addresses this gap by providing a population-specific, externally validated tool that achieves improved discrimination and more balanced risk stratification using routinely available clinical and laboratory data. Clinicians evaluating Korean patients with suspected coronary artery disease may benefit from incorporating K-CAD alongside existing guideline-endorsed models to support more individualized diagnostic decision-making. Future studies should perform formal recalibration of UDF and CAD2 to local disease prevalence, conduct decision curve analysis to quantify the net clinical benefit of K-CAD at specific threshold probabilities, and validate the model in broader East Asian and multiethnic cohorts to establish its wider applicability.

## BACKGROUND

Cardiovascular disease remains the foremost cause of morbidity and mortality worldwide, accounting for 17 million deaths annually.^1^ Coronary artery disease (CAD) contributes over 50% of this mortality, with increasing prevalence.^2^ The emergence of advanced data analytics and extensive datasets facilitates the development and validation of population-specific risk models for CAD. These models enhance decision-making for individualized diagnostic testing and strengthen patient–physician communication. By employing statistical learning methods, such tools can be integrated into comprehensive population strategies for cardiovascular prevention.^3^

The Updated Diamond–Forrester (UDF)^4^ and CAD consortium (CAD2)^5^ clinical models are pretest probability (PTP) tools endorsed by the American Heart Association/American College of Cardiology (AHA/ACC) and European Society of Cardiology (ESC) guidelines.^6–8^ UDF relies on clinical risk factors (CRFs) such as sex, age, and symptoms, and its predictive power for CAD is relatively low.^9^ CAD2, which incorporates additional medical history data including diabetes, hypertension, dyslipidemia, and smoking status and is constructed upon a high-quality dataset, exhibits a slightly improved predictive capability for CAD compared to UDF.^5,10^ Nevertheless, CAD2 remains relatively simplistic because it excludes routine laboratory blood tests such as lipid profiles, which are readily accessible and valuable for CAD prediction.

Importantly, these models may not be applicable to Asian populations, including Koreans, being predominantly based on Western demographic data. Previous research has demonstrated ethnic differences in predicting the presence or severity of CAD.^11^ Moreover, a distinct difference exists in the pattern of coronary atherosclerosis between Western and Korean adult populations.^12^ The “calibration hierarchy” concept, described by Van Calster and Vickers^13^, suggests that models developed in one setting often fail in another, not due to a lack of discrimination, but due to poor calibration-in-the-large (intercept) and calibration slope when applied to populations with different disease prevalence and case-mix. Western models applied to Asian cohorts frequently overestimate risk because they are not calibrated to the local baseline prevalence of obstructive disease.

Recently, statistical learning algorithms and penalized regression techniques, along with the inclusion of additional CRFs, have demonstrated promising results in predicting CAD.^14–16^ However, the external validation of these results and practical application of PTP models remain challenging, as most models yielding such promising outcomes are not publicly available or lack transparent reporting compliant with the TRIPOD guidelines.^17^ Therefore, the simplicity of UDF and CAD2 is a key feature that promotes their continued popularity in clinical practice.

We aimed to develop a novel PTP model for CAD, specifically tailored to the Korean population, by incorporating an expanded set of readily available CRFs, including routine laboratory results, such as lipid profiles. We proposed and comprehensively validated a CAD PTP model for Koreans (K-CAD) by employing two external datasets: outpatient CAD and nationwide health checkup data in Korea. Our validation included a comparison with the well-established UDF and CAD2 PTP models.

## METHODS

### Training Dataset and Data Harmonization

We developed a training dataset from the Korean population by employing data from three previously published randomized controlled trials (RCTs) and two retrospective cohorts (Table 1). The RCTs were CONSERVE (clinicaltrials.gov: NCT01810198), CREDENCE (clinicaltrials.gov: NCT02173275), and 3V FFR-FRIENDS (clinicaltrials.gov: NCT01621438), all of which were open-label, international, multicenter trials. The two retrospective cohorts comprised the PARADIGM registry (clinicaltrials.gov: NCT02803411) and Severance Coronary Computed Tomography Angiography (CCTA) registry.

A rigorous data harmonization protocol was implemented to ensure consistency across these heterogeneous sources. Variable definitions were standardized as follows: “Current Smoker” was defined as an active smoker within the last 30 days across all datasets. “Dyslipidemia” was defined as the current use of lipid-lowering medications or a diagnosis in the medical history, distinct from raw lipid values used as continuous predictors. Symptom categories were harmonized into “Typical Angina,” “Atypical Angina,” and “Non-cardiac Chest Pain” based on the Diamond–Forrester classification criteria recorded in each study’s case report forms. Laboratory values (lipid profile, creatinine level, and HbA1c level) were extracted from the baseline visit closest to the index imaging procedure.

The study was approved by the Institutional Review Board (IRB) of the Yonsei University College of Medicine (IRB Number: 4-2020-1314). All procedures adhered to the ethical standards of the Declaration of Helsinki, revised in 2013. Due to the retrospective nature of the study design, sample size calculations were unnecessary. Moreover, the requirement for informed consent was waived because participant selection was based on previously published anonymized data.

### External Validation Datasets

To assess the robustness of the K-CAD model irrespective of symptomatic status or risk level, two distinct external datasets were utilized. Initially, to validate the model in a high-risk symptomatic setting, data were retrospectively collected from the Division of Cardiology at Seoul National University Bundang Hospital (SNUBH). This local cohort (n=428), hereafter referred to as External Validation Cohort 1, with baseline characteristics detailed in Table 2, comprised symptomatic patients with suspected CAD who required invasive coronary angiography (ICA), representing a high-risk population in a tertiary care setting. For this cohort, the reference standard for CAD was defined as anatomically confirmed obstructive CAD (> 50% stenosis) via ICA.

Second, to evaluate generalizability in a low-risk general population, the National Health Insurance Service-Health Screening Cohort (NHIS-HEALS) dataset (2002–2013) (n=117,294), hereafter referred to as External Validation Cohort 2, was utilized. This dataset comprises individuals who underwent routine national health checkups. The requirement for informed consent was waived because the data was anonymized. Using the most recent dataset (2013), a cross-sectional analysis was conducted, excluding individuals with a history of heart disease, those without a baseline screening record, those who died before screening, and those with missing data. After these exclusions, 117,294 participants remained for the final analysis (Figure 1). In this asymptomatic screening cohort, the symptom variables (Typical Angina and Atypical Angina) were set as the reference category (Non-cardiac Chest Pain/Asymptomatic) for all participants, as they were undergoing routine health screening rather than symptom-driven evaluation.

**Figure 1.**
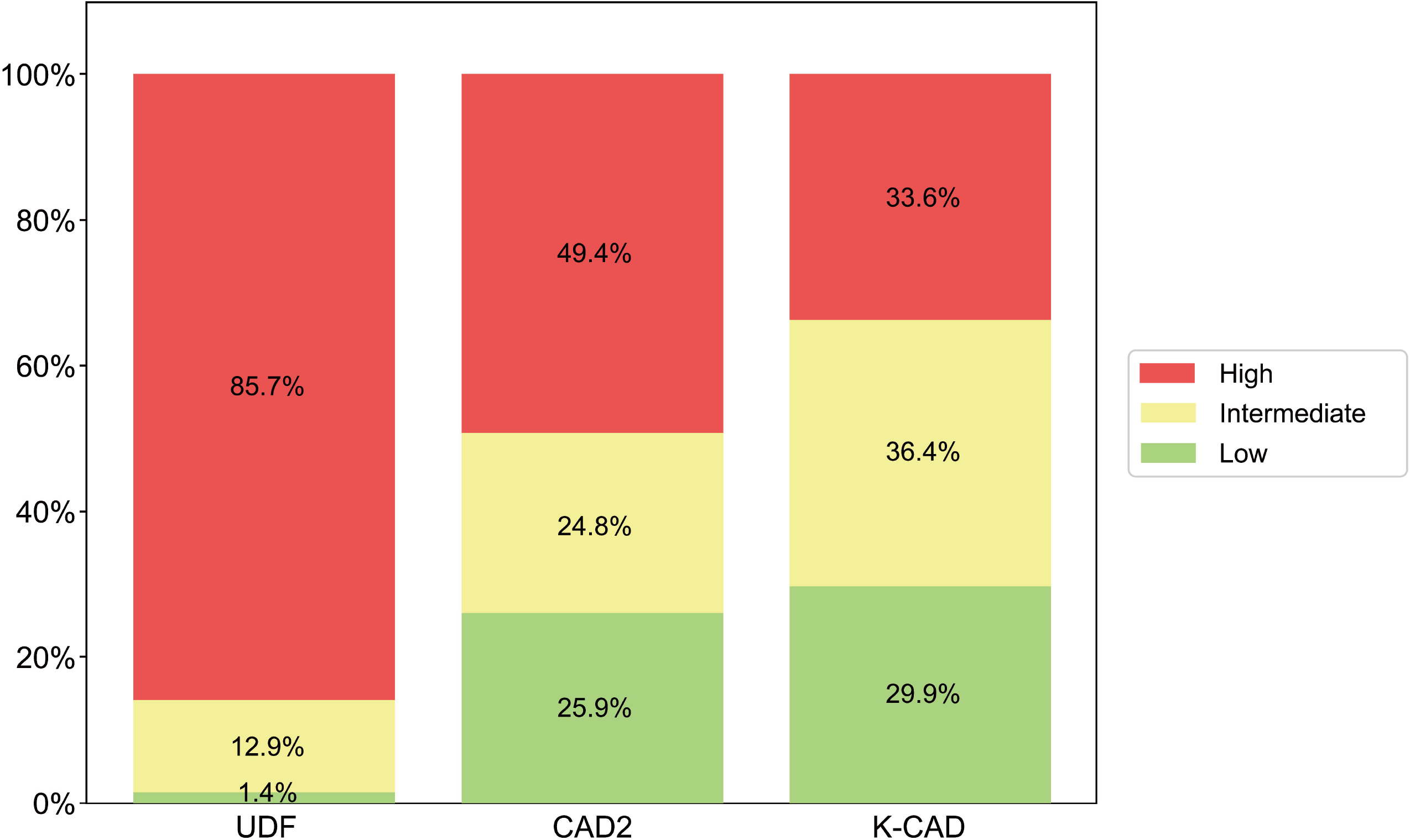
Data preparation flow. Data from five Korean cohorts (n=4,696) comprising patients that met all inclusion criteria were used as the training dataset. A local cohort from Bundang Seoul National University Hospital was also employed as an external validation set. Data from the NHIS-HEALS, excluding those for HbA1c and symptomatic variables but meeting all other criteria (n=117,294 patients), were used as an additional external validation set. Abbreviations: BMI, Body Mass Index; DM, Diabetes; HDL, High Density Lipoprotein Cholesterol; LDL, Low Density Lipoprotein Cholesterol; HbA1c, Hemoglobin A1c.

### Association with Clinical Angina Diagnosis (Surrogate Endpoint)

For External Validation Cohort 2 (NHIS-HEALS), the primary outcome was defined as the incident clinical diagnosis of angina (International Classification of Diseases 10th edition code: I20). This clinical surrogate endpoint was employed due to the impracticality and ethical concerns associated with routine anatomical evaluations of obstructive CAD in a nationwide screening cohort. A modified version of the K-CAD model, excluding glycated hemoglobin (HbA1c), was evaluated in this dataset, as detailed in the baseline characteristics (Supplementary Table 7). The findings from this cohort should be interpreted as an association between the model and clinical diagnosis, rather than anatomically confirmed obstructive CAD.

### CAD Definition

In this study, CAD was defined as obstructive CAD, characterized by > 50% diameter stenosis detectable via CCTA or ICA. The reference standard for CAD was ICA in the CONSERVE, CREDENCE, and 3V FFR-FRIENDS trials, while CCTA was used in the PARADIGM and Severance CCTA registries (Supplementary Table 6). Quantitative coronary angiography (QAngio XA®, Medis, The Netherlands) and semiautomated quantitative analysis (QAngio CT®, Medis, The Netherlands) were used to assess angiography and diameter stenosis in CCTA, respectively. For the External Validation Cohort 1 (SNUBH), ICA served as the reference standard for defining CAD.

### New Korean-CAD (K-CAD) Prediction Model Development

The new CAD prediction model for Koreans (K-CAD) incorporated all baseline clinical variables in Table 1 as predictors and utilized feature engineering techniques. Specifically, high-density lipoprotein (HDL) cholesterol and triglyceride levels were natural log-transformed to reduce skewness. Ages were discretized into four levels: 40–49 (Age_0), 50–59 (Age_1), 60–69 (Age_2), and ≥70 (Age_3) years. HbA1c levels were categorized into three groups: No Diabetes (reference), Diabetes with HbA1c 7–9% (HbA1c_1), and Diabetes with HbA1c ≥9% (HbA1c_2). Symptom categories were encoded using one-hot encoding, with “noncardiac chest pain serving as the reference category.

Ridge logistic regression (L2 regularization) was employed to identify the association between the predictors and CAD and to mitigate overfitting. The final probability of obstructive CAD was calculated as 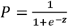, where ***Z*** is the linear combination of the intercept and the products of the coefficient and predictor values (Table 3). Moreover, the logistic model without HbA1c and the K-CAD version without HbA1c were validated using the NHIS-HEALS dataset.

### Statistical Analysis

We compared the descriptive statistics pertaining to the demographics, medical history, laboratory results, and symptoms of patients with and without obstructive CAD. Continuous variables are presented as means and standard deviations, whereas categorical variables are presented as counts and percentages. The Wilcoxon–Mann–Whitney nonparametric test was used to compare continuous variables, and categorical variables were compared using the chi-square test. Missing data was addressed through complete-case analysis for primary model development. Consequently, 540 patients with missing stratification variables or indeterminate stenosis were excluded from the detailed baseline comparison (Table 1) but contributed to the overall cohort assembly where feasible. A missingness audit was conducted to assess potential selection bias by comparing the characteristics of included and excluded patients.

We employed two distinct methodologies to evaluate the incremental predictive value of the K-CAD score over two established PTP models for CAD: the UDF and CAD2 clinical models. First, we generated receiver operating characteristic (ROC) curves and calculated the corresponding AUCs. Confidence intervals for sensitivity, specificity, and diagnostic accuracy were calculated using the Wilson scoring method. Subsequently, we performed ternary net reclassification improvement (NRI) analysis for low, intermediate, and high-risk categories based on risk percentages of <15%, 15 to <30%, and ≥30%, and calculated event-NRI (improvement in classifying cases) and nonevent NRI (improvement in classifying non-cases). The Python scikit-learn package was used to estimate the ridge regression coefficients and AUC.

### Calibration Analysis

To address potential calibration-in-the-large issues, in which Western models might overestimate risk in Asian populations due to differences in disease prevalence, we evaluated the need for recalibration. Recalibration-in-the-large involves adjusting the intercept of the logistic regression model while keeping the slope and other coefficients constant, thereby aligning the mean predicted risk with the observed event rate in the target population.^13^ We assessed calibration performance using the calibration intercept, calibration slope, and Brier score.

## RESULTS

### Data Collection and Study Population

A total of 4,696 Korean patients with suspected CAD who underwent ICA or CCTA from five previous studies (CONSERVE, CREDENCE, 3V FFR-FRIENDS, PARADIGM, and the Severance CCTA registry) were included in the raw training dataset. Of them, 4,156 patients had complete data for the primary analysis (Table 1), comprising 3,396 nonobstructive- and 760 with obstructive CAD. The remaining 540 patients were excluded from detailed baseline characteristic comparison due to missing values in specific stratification variables or indeterminate stenosis severity, although they contributed to the overall cohort assembly. A missingness audit comparing the 4,156 included patients with the 540 excluded patients revealed no significant differences in age or sex, although the excluded patients had higher rates of incomplete lipid profiles. For external validation, 428 patients from External Validation Cohort 1 (SNUBH) were included.

### Baseline Characteristics of Datasets

Patients with obstructive CAD exhibited a significantly higher burden of traditional cardiovascular risk factors compared to those without obstructive CAD across both cohorts (Tables 1 and 2). In both the training and external Validation Cohort 1, patients with obstructive CAD were significantly older, predominantly male, and had a higher prevalence of diabetes mellitus and hypertension. Although current smoking and dyslipidemia were significantly more common among patients with obstructive CAD in the training dataset, these differences were not statistically significant in the smaller validation cohort.

Compared with patients without obstructive CAD, those with obstructive CAD were more likely to have lower low-density lipoprotein (LDL) cholesterol (training data, 108·4 vs. 114·9, p<0·001; validation data, 87·0 vs. 91·3, p=0·17), lower HDL cholesterol (training data, 46·1 vs. 50·5, p<0·001; validation data, 47·6 vs. 54·7, p<0·001), and higher HbA1c (training data, 6·00 vs. 5·89, p<0·001; validation data, 6·27 vs. 5·92, p<0·001) levels. Additionally, compared with patients without obstructive CAD, those with obstructive CAD were more likely to have typical angina symptoms (training data, 31·1% vs. 9·3%; validation data, 52·8% vs. 29·0%) (Tables 1 and 2). Supplementary Table 7 presents the baseline characteristics of the nationwide validation dataset used for the risk factor model.

### Estimates for CAD Prediction Obtained from Ridge-Regression

The estimates from the ridge-regression model indicate that male sex (odds ratio [OR] 2·25, 95% confidence interval [CI] 1·89–2·67), hypertension (OR 1·22, 95% CI 1·04–1·45), dyslipidemia (OR 1·60, 95% CI 1·37–1·88), and typical angina (OR 2·60, 95% CI 2·14–3·15) exhibit a significant positive association with obstructive CAD. For discretized age and HbA1c levels, increasing levels correlated with larger regression coefficient estimates (from 0·5157 to 2·007 for age and from 0·2123 to 0·3953 for HbA1c). Moreover, their effective size similarly increased (from OR 1·67 to 7·44 for age and from OR 1·24 to 1·48 for HbA1c). In contrast, body mass index (BMI) and HDL cholesterol levels were significantly negatively associated with obstructive CAD (Table 3).

### Model Performances with Continuous ROC and NRI Analysis: K-CAD vs. UDF vs. CAD2

Continuous ROC analysis yielded the following AUC values: 0·68 (95% CI 0·63–0·73) for UDF, 0·71 (95% CI 0·67–0·76) for CAD2, and 0·76 (95% CI 0·71–0·80) for K-CAD. K-CAD exhibited a significantly higher AUC than UDF and CAD2 (p <0·001 for UDF and *p*<0·05 for CAD2) (Figure 2). The diagnostic accuracy of K-CAD was 70·8% (95% CI 67·3–75·5); sensitivity 80·4% (95% CI 74·6–85·2); specificity 61·2% (95% CI 54·5–67·5); positive predictive value 67·5% (95% CI 63·2-81·8); and negative predictive value 75·7% (95% CI 63·3–81·8). This was superior to UDF, which had an accuracy of 65·2% (95% CI 61·2–69·9), and CAD2, with an accuracy of 67·5% (95% CI 63·8 –72·2).Model calibration based on observed and predicted risk indicated that while K-CAD fit the training set well, it slightly underestimated the predicted risk compared to the observed risk in the high-risk group of External Validation Cohort 1.

**Figure 2.**
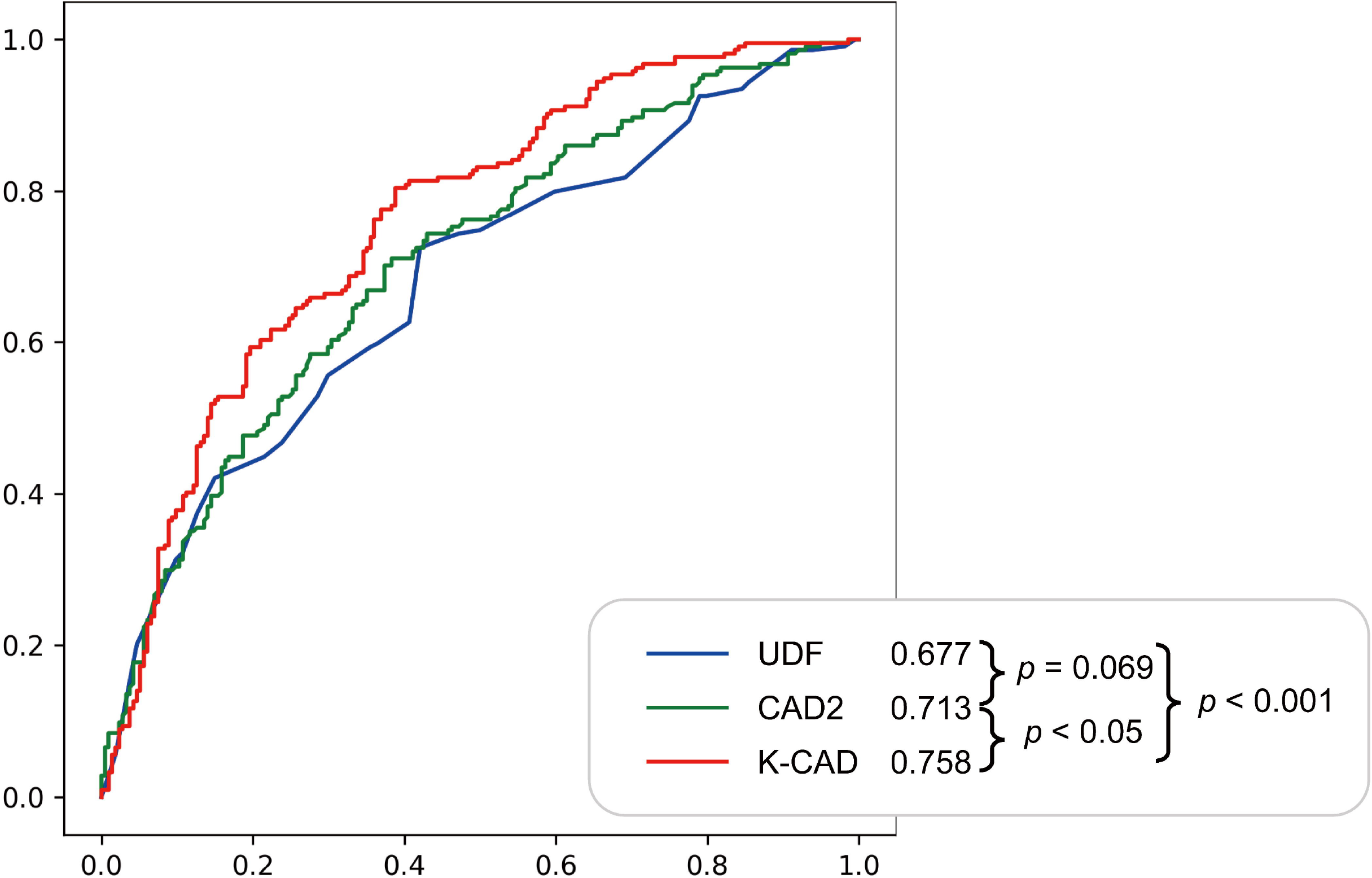
Continuous ROC analysis for three CAD prediction models: UDF (blue), CAD2 (green), and K-CAD (red). The evaluated area under the ROC curve was 0·677 for UDF, 0·713 for CAD2, and 0·758 for K-CAD. K-CAD exhibited significantly higher AUC than UDF and CAD2 (p < 0·001 for UDF and p < 0·05 for CAD2). However, no significant differences were observed between UDF and CAD2 (p = 0·069). Abbreviations: ROC, Receiver Operating Characteristic; CAD, Coronary Artery Disease; UDF, Updated Diamond– Forrester score; CAD2, CAD Consortium; K-CAD, CAD PTP model for Koreans (our CAD prediction model).

When the UDF model was utilized, only 1·4% of the patients were deemed low-risk, whereas CAD2 increased this proportion to 24·8%, and K-CAD elevated it to 36·4%. Conversely, the UDF model classified 85·7% of patients as high-risk, with this proportion decreasing to 49·3% with CAD2 and 33·6% with K-CAD (Figure 3).

**Figure 3.**
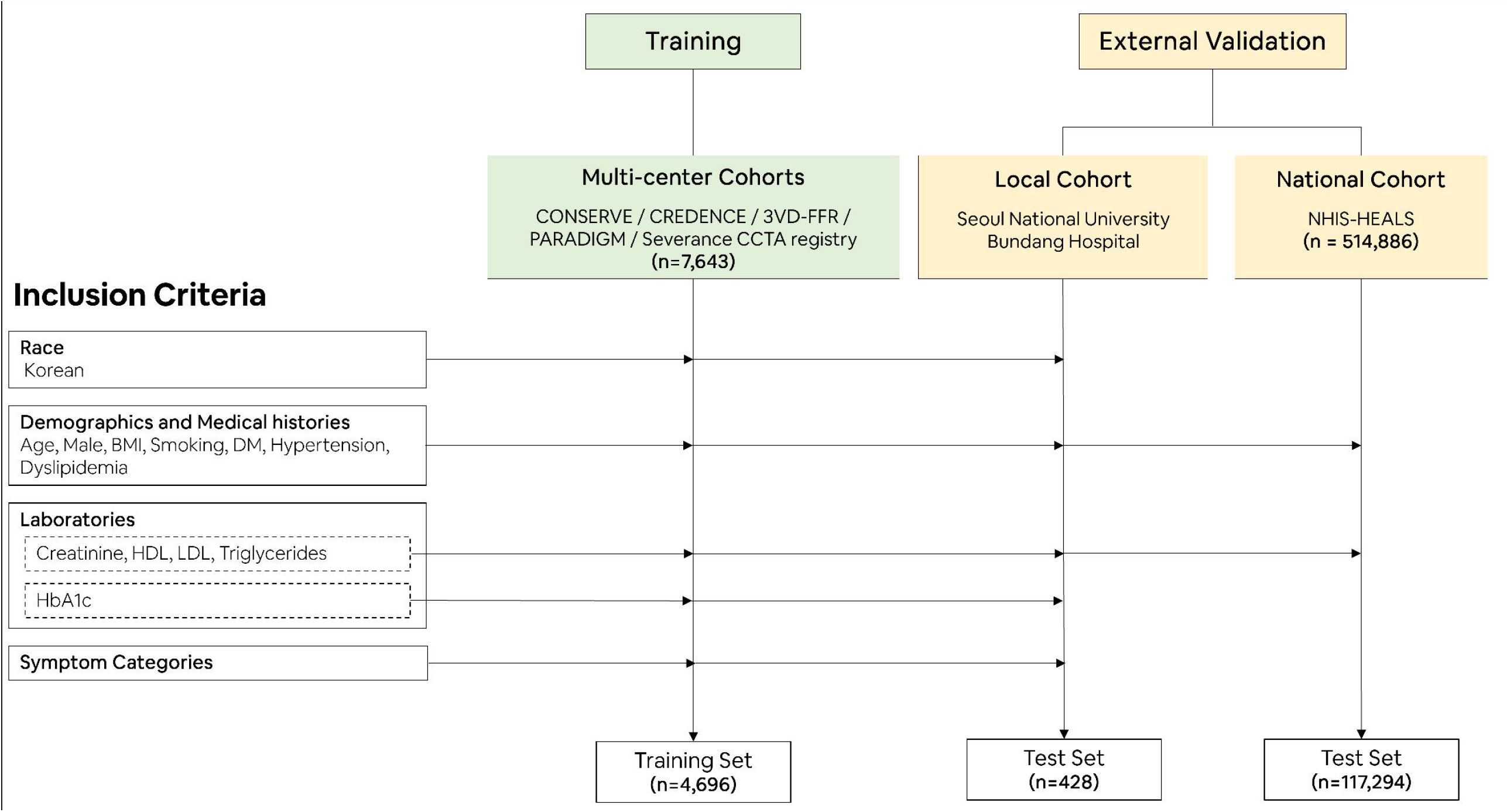
Distribution of the population in low (green), intermediate (yellow), or high (red) pretest probability scores for the UDF, CAD2, and K-CAD. The distribution of predicted risk is shown for patients categorized into three risk groups: low, intermediate, and high-risk. UDF model predicted 1·4% of patients to be low-risk, 12·9% to intermediate-risk, and 85·7% to high-risk. CAD2 model predicted 25·9% of patients to be low-risk, 24·8% to intermediate-risk, and 49·3% to high-risk. K-CAD model predicted 29·9% of patients to be low-risk, 36·4% to intermediate-risk, and 33·6% to high-risk. Abbreviations: UDF, Updated Diamond–Forrester score; CAD2, CAD Consortium; K-CAD, CAD PTP model for Koreans (our CAD prediction model).

An NRI analysis was performed to assess the reclassification performance. Among patients without obstructive CAD (non-events), K-CAD correctly reclassified a significant proportion into lower-risk categories compared to UDF and CAD2 (Non-event NRI). Specifically, 79·9% (171/214) of non-obstructive patients classified as high-risk by UDF were reclassified as lower risk by K-CAD. However, among patients with obstructive CAD (events), K-CAD reclassified some patients into lower-risk categories, indicating a tradeoff in which improved specificity (identifying non-cases) was accompanied by a reduction in sensitivity (Event NRI). The overall NRI values for the K-CAD score compared with those of UDF and CAD2 were 0·355 and 0·121, respectively (Supplementary Table 8).

Additionally, the generalizability of the K-CAD PTP model was tested in a nationwide health check-up population (External Validation Cohort 2). Consistent with previous results, the K-CAD model outperformed the other models on this dataset. Continuous ROC analysis yielded AUC estimates of 0·61 (95% CI 0·58–0·64) for UDF, 0·66 (95% CI 0·63–0·69) for CAD2, and 0·67 (95% CI 0·64–0·70) for the K-CAD version without HbA1c. As indicated by the standard errors, K-CAD and CAD2 demonstrated significantly higher AUC estimates than UDF, and K-CAD demonstrated slightly higher AUC estimates than CAD2, although these were not statistically significant. The K-CAD score calculator, featuring the estimated model parameters, is available online at https://metaeyes.io/med_scores/k_cad.

## DISCUSSION

In this study, we employed ridge-penalized logistic regression to develop an advanced PTP model, K-CAD, optimized specifically for the Korean population and constructed using a high-quality database. The K-CAD score incorporates readily available routine blood test results, which are beneficial for predicting CAD, alongside basic CRFs. This refined CAD score exhibited superior performance compared to those of both UDF and CAD2 when validated using a dual-track strategy comprising a high-risk Korean outpatient clinic cohort and a large nationwide health checkup population dataset.

Previous population-based studies in Korea have highlighted the suboptimal CAD prediction performance of UDF and CAD2 models. In one study, patients with acute chest pain visiting the emergency department had AUCs of 0·72 for UDF and 0·74 for CAD2.^19^ Similarly, among patients hospitalized for ICA with angina, the AUCs were 0·69 for UDF and 0·72 for CAD2.^9^ Our study yielded consistent results, reporting AUCs of 0·68 for UDF and 0·71 for CAD2. Additionally, the CAD2 model’s performance was reported to be insufficient in a mixed Asian cohort study, with an AUC of 0·72.^20^ These findings reinforce our assertion that the UDF and CAD2 models are not optimal for East Asians or specifically, Korean demographics.

Such discrepancies may be attributable to the “calibration hierarchy” described by Van Calster and Vickers.^13^ Models developed in Western populations (UDF, CAD2) often fail in Asian populations not necessarily due to poor discrimination (ranking of patients), but rather, due to poor calibration-in-the-large (intercept) and calibration slope. The baseline prevalence of obstructive CAD and the weight of specific risk factors differ considerably between Western and Korean populations.^11,12^ By training the K-CAD specifically on Korean data, we effectively addressed these calibration issues, creating a PTP model that aligns with the unique requirements of the Korean population. It is important to note that the superior performance of K-CAD may be partly driven by this prevalence adjustment (intercept), rather than by better discrimination. Future studies should perform formal recalibration-in-the-large of the UDF and CAD2 on the local prevalence to isolate the discriminative value of the additional predictors included in K-CAD.

Developing risk-scoring systems from clinical data frequently addresses the challenge of overfitting. To counter this, we implemented a ridge estimator, which is a shrinkage-based method, for parameter estimation. By leveraging this technique on expansive and balanced clinical data from various cohort studies, the K-CAD score slightly reduced overfitting, attesting to its high accuracy during external validation. While recent studies have explored advanced machine learning models such as XGBoost or ensemble methods for CAD prediction,^16,21^ we selected ridge-penalized logistic regression to maintain model interpretability and facilitate ease of clinical implementation, while still benefiting from regularization to handle multicollinearity among risk factors. Therefore, the K-CAD is poised to become an ideal PTP model for CAD, potentially benefiting a diverse range of Koreans and possibly East Asian individuals across different cardiovascular risk tiers.

Our analysis identified associations between risk factors and obstructive CAD, consistent with the findings of previous studies. Factors such as male sex, hypertension, dyslipidemia, typical angina, creatinine level, age, and HbA1c level were positively associated with obstructive CAD. Conversely, BMI and HDL cholesterol levels display an inverse relationship with obstructive CAD.^2,10,16^ Notably, our baseline data showed lower LDL cholesterol levels in the obstructive CAD group. This inverse association possibly reflects confounding by indication, resulting from the inclusion of patients who were already on baseline lipid-lowering therapies (e.g., statins), particularly within the large-scale CCTA registries comprising our training dataset. In our study, atypical angina was inversely associated with obstructive CAD, aligning with the findings of one of the most extensive database studies and a recent mixed Asian cohort^20,22^ but contrasts with the UDF and CAD2 score models. This discrepancy may arise from the vague nature of atypical chest pain, which sometimes overlaps with sources of non-cardiac pain, such as gastrointestinal ailments. Consequently, the recent AHA/ACC guidelines caution against relying on atypical chest pain as a diagnostic indicator, highlighting the urgency for advancements in PTP models tailored to this ambiguous patient segment.

While the National Institute for Health and Care Excellence guidelines^17^ differ, the current AHA/ACC and ESC guidelines still advocate stratifying the PTP for CAD. Nevertheless, our findings suggest that the existing UDF and CAD2 models favor the high-risk group, inadequately representing low- and intermediate-risk categories, proposed by previous research.^16,19,20^ In contrast, the K-CAD model effectively repositioned a considerable portion of the high-risk demographic into low- and intermediate-risk categories, thereby refining risk stratification. Such recalibration may deter unnecessary downstream testing. Notably, UDF and CAD2 have maintained popularity due to their user-friendly designs. In contrast, K-CAD, while preserving practicality, elevates accuracy by integrating additional laboratory metrics such as HbA1c, creatinine, and lipid panels, which are generally readily available. Therefore, although K-CAD was tailored to the Korean demographic, its enhanced design could position it as an advanced PTP model applicable to diverse ethnicities.

In a recent investigation, machine-learning-based PTP models demonstrated a notable improvement, achieving an AUC of approximately 0·8 within a Korean demographic, underscoring the effectiveness of machine learning techniques.^21^ Nevertheless, these models were not validated using an external database, and the model parameters were not disclosed publicly, thereby limiting reproducibility.^16^ Consequently, these findings encountered skepticism because of the potential overfitting risks inherent in machine learning models. Moreover, that study identified troponin as the primary determinant for predicting CAD, suggesting that the model may be inappropriate for patients presenting with stable chest pain. Conversely, the K-CAD model not only exhibited strong performance in external validation but also introduced an online K-CAD score calculator complete with the model’s parameters. Recent comparative studies such as the IJC-CAD/ML-CAD2 analysis^21^ and COME-CCT-PTP evaluation^20^ have demonstrated that machine-learning-enhanced models can achieve AUCs of up to 0·86. Our K-CAD model, using a simpler ridge regression approach for interpretability, aligns with the trend of improving conventional models through improved local calibration and feature integration.

Our study has certain limitations. First, as our focus was on developing a CAD prediction model tailored for domestic use, its applicability to non-Korean populations may be limited. Second, we did not exclude asymptomatic patients from our study to develop a model applicable to a wider range of cases, whereas the UDF and CAD2 models were developed solely for symptomatic patients. Therefore, when clinicians utilize our K-CAD model to diagnose CAD in symptomatic patients, it is highly recommended to refer to the results from the UDF and CAD2 models for more comprehensive decision-making.

Third, the definitions and reference standards for the CAD outcomes inherently varied across our datasets, introducing labeling heterogeneity. In the training phase, we utilized a mixed reference standard that combined ICA and CCTA. Because CCTA tends to overestimate stenosis severity compared with the gold-standard ICA, this could introduce a labeling bias. Nevertheless, the stability of the model was corroborated by its robust predictive performance in the External Validation Cohort 1 (SNUBH), which relied solely on ICA.

Fourth, for the asymptomatic NHIS-HEALS validation cohort (External Validation Cohort 2), we relied on the clinical diagnosis of angina (ICD-10 I20) as a surrogate outcome rather than anatomically established obstructive CAD. We acknowledge that an ICD-based clinical diagnosis differs considerably from an anatomically confirmed CAD diagnosis. However, routine anatomical evaluations (ICA or CCTA) in a nationwide primary prevention screening cohort of over 100,000 asymptomatic individuals are neither feasible nor ethical. Therefore, while we recognize this methodological limitation, utilizing this surrogate endpoint was a feasible approach for evaluating the generalizability of the model in a large-scale, real-world screening setting. The results in this cohort should be interpreted as an association with clinical angina diagnosis rather than direct validation of obstructive CAD prediction.

Fifth, in our comparison of K-CAD with UDF and CAD2, we did not recalibrate the competitor models to align with the local prevalence in the current analysis. It is plausible that the superior performance of K-CAD is partly attributable to better calibration of local disease prevalence rather than solely better discrimination. Future research should explore whether recalibrating the UDF/CAD2 intercepts reduces the performance disparity.

Finally, a model was developed using the heterogeneous population dataset from each trial. This diversity enabled the inclusion of a wide range of risk groups, from low to high, which is an advantage compared to traditional models that often exhibit bias towards certain risk groups. However, our model did not incorporate various factors associated with obstructive CAD such as supplemental laboratory results, CAC scores, or echocardiographic parameters, all of which could potentially augment performance. Although we achieved substantial improvements by utilizing commonly available clinical data, further research is required to develop an even more precise PTP model in a cost-effective manner, without relying on expensive downstream modalities. Such improvements can be realized by leveraging advanced deep learning techniques.

## CONCLUSIONS

We developed a general risk factor-based CAD prediction model using a large-scale, well-balanced database, and externally validated its reliability. This ridge regression-based online K-CAD calculator may be beneficial in the clinical decision-making process for patients with suspected CAD who visit outpatient clinics.

## Data Availability

The training dataset was derived from previously published clinical trials and registries (CONSERVE, CREDENCE, 3V FFR-FRIENDS, PARADIGM, and Severance CCTA registry), and access is subject to the data sharing policies of each original study. The External Validation Cohort 1 (SNUBH) data are available upon reasonable request to the corresponding author. The External Validation Cohort 2 (NHIS-HEALS) data are available through the National Health Insurance Service of Korea (https://nhiss.nhis.or.kr) upon approval of a data use application. The K-CAD model parameters are publicly available in Table 3 of the manuscript, and the online calculator is accessible at https://metaeyes.io/med_scores/k_cad.

## DECLARATIONS

### Ethics approval and consent to participate

This study was approved by the Institutional Review Board of Yonsei University College of Medicine (IRB Number: 4-2020-1314). Owing to the retrospective nature of the study design, the requirement for patient informed consent was waived, as participant selection was based on previously published anonymized data.

### Consent for publication

Not applicable.

### Data Availability Statement

The data underlying this article cannot be shared publicly because of institutional regulations and privacy concerns of the individuals who participated in the study. The data will be shared upon reasonable request from the corresponding authors.

### Authors’ contributions

**HBP:** Conceptualization, Methodology, Investigation, Data curation. **JL:** Conceptualization, Visualization, Methodology, Investigation, Data curation. **YEY:** Methodology, Investigation, Data curation. **YH:** Conceptualization, Methodology, Investigation, Data curation, and supervision. **WK:** Conceptualization, Visualization, Methodology, Investigation, Data curation, and supervision. **GK:** Data curation and formal analysis. **SK:** Data curation and formal analysis. **BS:** Methodology, Software. **TP:** Methodology, Software. **JJ:** Methodology, Software. **HJ:** Methodology, Software. **JMS:** Data curation and Supervision **WK:** Methodology, Supervision **HJC:** Data curation, methodology, and supervision. All authors participated in the drafting process and conducted a thorough critical review. Subsequently, all authors approved the final version to be published.

## Acknowledgments

Not applicable

