## Supplementary Materials for "Machine learning-based advanced coronary artery disease pretest probability model: Comparison with conventional pretest probability models"

**Supplementary Table 1. CONSERVE inclusion and exclusion criteria**

**Inclusion criteria**

1) Moderate or severe angina, which improves with mild with medical therapy
2) Mild or moderate angina intolerant to medical therapy
3) Any angina, not evaluable by noninvasive stress testing
4) Heart failure with normal ejection fraction of unknown etiology
5) Symptomatic (chest pain) + abnormal NIST
6) Asymptomatic + 2 RF + abnormal NIST
7) Worsening NIST
8) Recurrent hospitalization for chest pain + abnormal/equivocal NIST
9) Low-risk surgery, stable angina
10) Low-risk surgery, medically stabilized moderate or severe stable angina
11) High-risk surgery with equivocal NIST
12) Asymptomatic, high-risk occupation
13) Vascular surgery with >2 RF
14) Perioperative MI
15) High risk for coronary disease when other cardiac surgical procedures are planned (e.g., pericardiectomy or removal of chronic pulmonary emboli)
16) Prospective immediate cardiac transplant donors whose risk profile increases the likelihood of coronary disease
17) Asymptomatic patients with Kawasaki disease who have coronary artery aneurysms on echocardiography
18) Before surgery for aortic aneurysm/dissection in patients without known coronary disease
19) Recent blunt chest trauma and suspicion of acute MI, without evidence of preexisting CAD

**Exclusion criteria**

1) Known CAD (past MI, PCI, CABG, or diagnosed by cardiac catheterization without intervention)
2) Acute coronary syndrome or MI at time of enrollment
3) Planned intervention or bypass surgery
4) Known complex congenital heart disease
5) Planned invasive angiography for reasons other than CAD
6) Noncardiac illness with life expectancy <2 years
7) Inability to provide written informed consent
8) Concomitant participation in another clinical trial in which patient is subject to investigational drug or device
9) Pregnant women
10) Allergy to iodinated contrast agent
11) Serum creatinine >1·5 mg/dL or GFR <30 mL/min
12) Baseline irregular heart rhythm
13) Heart rate ≥100 beats/min
14) Systolic blood pressure ≤90 mm Hg
15) Contraindications to β-blockers or nitroglycerin
16) Body mass index >35 kg/m^2^
17) Known complex congenital heart disease
18) Age <18 years

NIST, National Institute of Standards; RF, risk factor; MI, myocardial infarction; CABG, coronary artery bypass graft; GFR, glomerular filtration rate.

**Supplementary Table 2. CREDENCE inclusion and exclusion criteria**

**Inclusion criteria**

1) Age ≥18 years
2) Patients scheduled to undergo clinically indicated non-emergent invasive coronary angiography

**Exclusion criteria**

1) Known coronary artery disease (myocardial infarction, percutaneous coronary interventions, and coronary artery bypass graft)
2) Hemodynamic instability
3) Inability to provide written informed consent
4) Concomitant participation in another clinical trial in which an individual is subject to investigational drug or device
5) Pregnancy or unknown pregnancy status
6) Absolute contraindication to iodinated contrast due to prior near-fatal anaphylactoid reaction (laryngospasm, bronchospasm, cardiorespiratory collapse, or equivalent)
7) Impaired chronic renal function (serum creatinine ≥1·7 mg/dL or glomerular filtration rate <30 mL/min)
8) Baseline irregular heart rhythm (such as atrial fibrillation)
9) Heart rate ≥100 beats/min
10) Systolic blood pressure ≤90 mm Hg
11) Contraindications to β blockers or nitroglycerin or adenosine

**Supplementary Table 3. 3V FFR-FRIENDS inclusion and exclusion criteria**

**Inclusion criteria**

1) Patient must be at least 18 years of age.
2) Patient must have stenosis (>30% by visual estimate) in all 3-epicardial coronary arteries.
3) FFR should be measured at all 3-vessels at the end of a procedure.

**Exclusion criteria**

1) Depressed left ventricular systolic function (ejection fraction <35%)
2) ST-elevation myocardial infarction within 72 h
3) Prior coronary artery bypass graft surgery
4) Creatinine level ≥2·0 mg/dL or dependence on dialysis
5) Abnormal final myocardial flow (TIMI flow <3)
6) Planned bypass surgery
7) Failed FFR measurement
8) Failed intended revascularization

Abbreviations: FFR, fractional flow reverse; TIMI, thrombolysis in myocardial infarction.

**Supplementary Table 4. PARADIGM inclusion and exclusion criteria**

**Inclusion criteria**

1) Patients who underwent two or more clinically indicated CCTA with 64-detector rows or greater for CAD evaluation.
2) At least a 2-year interval between the baseline and follow-up CCTAs
3) Prospective data collection for CAD risk factors

**Exclusion criteria**

1) Unavailable clinical or laboratory data within 1 month from baseline CCTA or follow-up CCTA
2) Uninterpretable CCTA

Abbreviations: CCTA, coronary computed tomography angiography; CAD, coronary artery disease.

**Supplementary Table 5. Severance CCTA registry inclusion and exclusion criteria**

**Inclusion criteria**

1) Consecutive patients who underwent both coronary CTA and XECG within 90 days for evaluation of suspected CAD at Severance Cardiovascular Hospital from May 2003 to April 2009 without any other cardiovascular testing

**Exclusion criteria**

1) Age <30 years
2) History of prior myocardial infarction, coronary revascularization, or cardiac transplantation
3) Inadequate XECG
4) Insufficient medical records or uninterpretable coronary CTA results
5) Without at least 1 of following symptoms or signs: angina, angina equivalent symptoms, or abnormal resting electrocardiography results

Abbreviations: CTA, computed tomography angiography; CAD, coronary artery disease; XECG, exercise electrocardiography.

**Supplementary Table 6. Datasets associated with the study**

|  | **Training**  (n=4,696) | **Test**  (n=428) | **Nationwide Validation**  (n=117,294) |
| --- | --- | --- | --- |
| **Data Source** | CONSERVE (n=878) | Seoul National University Bundang Hospital |  |
|  | CREDENCE (n=350) |  |  |
|  | 3VD FFR-FRIENDS (n=145) |  | NHIS-HEALS |
|  | PARADIGM (n=902) |  |  |
|  | Severance CCTA registry (n=2,421) |  |  |
| **Nation** | Korean | | |
| **Patient Characteristics** |  | | |
| Demographics | Age, Male, BMI, Smoking | | |
| Medical History | DM, Hypertension, Dyslipidemia | | |
| Laboratory results | Creatinine, HDL, LDL, Triglycerides | | |
|  | HbA1c | |  |
| Symptom categories | Typical angina, atypical angina, non-cardiac | |  |
| **Labeling for CAD** | 50% DS by CCTA & ICA | 50% DS by ICA | Angina (I20) in ICD-10 |

Abbreviations: BMI, body mass index; DM, Diabetes mellitus; CCTA, coronary computed tomography angiography; ICA, invasive coronary angiography; DS, diameter stenosis; NHIS-HEALS, National Health Insurance Service-Health Screening Cohort; ICD-10, International Classification of Diseases 10th edition; CAD, coronary artery disease; HbA1c, glycated hemoglobin; HDL, high-density lipoprotein; LDL, low-density lipoprotein.

**Supplementary Table 7.** **Baseline characteristics of the nationwide validation dataset for the risk factor model**

|  | **Total**  (n=117,294) | **Non-obstructive CAD**  (n=116,910) | **Obstructive CAD**  (n=384) | p-value |
| --- | --- | --- | --- | --- |
| **Demographics and Medical history** | |  |  |  |
| Age, years | 60·10 ± 7·92 | 60·18 ± 7·91 | 66·08 ± 9·46 | <·001 |
| Male, n (%) | 67,199 (57·29) | 66,961 (57·28) | 238 (61·98) | <·001 |
| Body mass index, kg/m^2^ | 23·97 ± 2·90 | 23·97 ± 2·90 | 24·12 ± 2·82 | 0·29 |
| Current Smoker, n (%) | 19,543 (16·66) | 19,488 (16·67) | 55 (14·32) | 0·21 |
| Diabetes mellitus, n (%) | 11,630 (9·92) | 11,545 (9·88) | 85 (22·14) | <·001 |
| Hypertension, n (%) | 34,418 (29·34) | 34,237 (29·28) | 181 (47·14) | <·001 |
| Dyslipidemia, n (%) | 7,083 (6·04) | 7,058 (6·04) | 25 (6·51) | 0·70 |
| **Laboratory data** | |  |  |  |
| Creatinine, mg/dL | 0·92 ± 0·47 | 0·92 ± 0·47 | 0·99 ± 0·66 | 0·0015 |
| HDL cholesterol, mg/dL | 53·60 ± 15·81 | 53·61 ± 15·82 | 50·05 ± 12·32 | <·001 |
| LDL cholesterol, mg/dL | 118·96 ± 34·53 | 118·96 ± 34·54 | 119·04 ± 34·33 | 0·96 |
| Triglycerides, mg/dL | 133·83 ± 83·26 | 133·81 ± 83·28 | 140·21 ± 77·54 | 0·13 |

Abbreviations: Continuous values are presented as means ± SDs, and categorical values are presented as n (%)

CAD, coronary artery disease; LDL, low-density lipoprotein; HDL, high-density lipoprotein.

**Supplementary Table 8. Reclassification of study participants into groups with low, medium, and high risk of CAD for U-DFS, CAD2, and K-CAD risk scores**

|  |  | Non-obstructive CAD (n=214) | | | |  | Obstructive CAD (n=214) | | | |
| --- | --- | --- | --- | --- | --- | --- | --- | --- | --- | --- |
|  |  | K-CAD | | | |  | K-CAD | | | |
|  |  | Low | Medium | High | Total |  | Low | Medium | High | Total |
| **U-DFS** | | | | | | | | | | |
|  | Low | 4 | 0 | 0 | 4 |  | 2 | 0 | 0 | 2 |
|  | Medium | 34^C^ | 7 | 0 | 41 |  | 9^I^ | 5 | 0 | 14 |
|  | High | 57 ^C^ | 80 ^C^ | 32 | 169 |  | 22^I^ | 64^I^ | 112 | 198 |
| **CAD2** | | | | | | | | | | |
|  | Low | 67 | 15^I^ | 0 | 76 |  | 18 | 15^C^ | 1^C^ | 34 |
|  | Medium | 27^C^ | 38 | 1^I^ | 61 |  | 12^I^ | 23 | 5^C^ | 40 |
|  | High | 6^C^ | 34^C^ | 31 | 77 |  | 3^I^ | 31^I^ | 106 | 140 |
| Total | | 95 | 87 | 32 |  |  | 33 | 69 | 112 |  |

U-DFS, Updated Diamond–Forrester; CAD 2, CAD Consortium clinical; C, correctly reclassified; I, incorrectly reclassified.

NRI for patients without obstructive CAD: Compared to U-DFS: 171/214=0·799; Compared to CAD2:51/214=0·238

NRI for patients with obstructive CAD: Compared to U-DFS: -95/214=-0·444, Compared to CAD2: -25/214=-0·117
